# Variations in iron profile among dialysis adherent chronic kidney disease patients and compare with the non-adherence chronic kidney disease patients

**DOI:** 10.1101/2025.01.03.25319943

**Authors:** Collince Odiwuor Ogolla, Lucy W. Karani, Stanslaus Musyoki

**Author notes:** **Corresponding Author:** Collince Odiwuor Ogolla.

## Abstract

**Background:** Chronic kidney disease (CKD) constitutes one of the most important global health challenges and iron deficiency (ID) anemia are both frequent complications, especially in patients on dialysis. Dialysis treatment is an important aspect of addressing these complications, yet the treatment adherence rate is low in CKD patients and the association between dialysis adherence and differences in iron profiles among CKD patients is unclear.

**Objectives:** This study aimed to assess differences in iron profile in dialysis-adherent and non-adherent chronic kidney disease subjects and analyze the profiles between both groups.

**Methods:** One hundred twenty patients undergoing hemodialysis were included in this cross-sectional study, divided into two groups based on two subcategories of dialysis adherence-adherent (n=60) and nonadherent (n=60). The parameters of iron profile-also defined as serum ferritin, transferrin saturation (TSAT), hemoglobin, and serum iron-were studied. Further, multivariate regression analysis was carried out, adjusting possible confounders such as age, sex, diabetes, and duration of dialysis. Results: Serum ferritin was significantly higher among patients in the adherent group when compared with those in the non-adherent group (235.6 120.2 ng/mL vs. 185.2 105.3 ng/mL; p=0.03), TSAT (33.4 9.3% vs. 28.8 10.2%; p=0.02), and hemoglobin level (11.5 1.8 g/dL vs. 10.2 2.1 g/dL; p=0.04). That is, non-adherence to the therapy was associated with a significantly higher number of patients having iron deficiency anemia (63% in non-adherent vs. 40% in adherent patients; p=0.01). Multivariate analysis confirmed that dialysis adherence was independently associated with better iron status (p<0.05 for all parameters).

**Conclusion:** In hemodialysis patients, adherence to dialysis presents as a strong predictor of better iron profile. More “iron parameters” were better and showed a decreased reliance on iron deficiency anemia for adherents. Such improvement, however, may significantly reduce anemia-associated complications through strategies that improve adherence to dialysis treatment by optimizing iron metabolism among chronic kidney disease patients.

**What this study adds:** This study demonstrates a significant relationship between adherence to dialysis and improved iron profiles, showing that a chronic kidney disease patient who is adherent has a better iron status and a reduced rate of having iron deficiency anemia compared with an individual who is nonadherent.

## Introduction

Chronic kidney disease (CKD) has become one of the major world health problems, affecting an estimated 10-15% of the adult population globally [1]. Recurrent dialysis is a lifesaving lifesaving requirement as CKD progresses to end-stage renal disease (ESRD). Among the many forms of dialysis available for ESRD management is hemodialysis, which focuses on metabolic waste and fluid imbalance. Notably, patients subjected to hemodialysis often experience iron deficiency and anemia due to reduced erythropoiesis, blood loss during dialysis, and impaired iron utilization [2, 3]

Iron deficiency in chronic kidney disease (CKD) is one of the most important causes of anemia, which results in an increase in fatigue, decreased quality of life, and possibly poorer clinical outcomes, with increased morbidity and mortality [4, 5]. Iron therapy, oral iron supplementation, or intravenous iron is employed most often to improve patients’ iron status and manage their anemia. However, it has become difficult to achieve an ideal iron balance for patients, especially when they do not adhere sufficiently to dialysis treatment.

This factor makes adherence to dialysis a crucial factor in the effective management of chronic kidney disease and its associated complications. Reports have indicated that poor adherence to dialysis can manifestly worsen the outcomes that would otherwise be more favorable such as inadequate dialysis, fluid overload, and malnutrition [6, 7]. In terms of anemia management, non-compliance may result in decreased iron stores which may in turn affect hemoglobin levels and transferrin saturation (TSAT) [8]. The association between adherence to dialysis and clinical outcomes has been clearly established, yet the specific effects of adherence on the differences in iron profile of CKD patients remain unknown.

## Methodology

This study utilized an observational cross-sectional design that aimed to examine the differences in iron profile among dialysis adherent vs. nonadherent chronic kidney disease (CKD) patients. The study was approved by the institutional review board (IRB). All consenting parties signed consent forms as a requirement to fulfill the ethical requirements of the Declaration of Helsinki.

The study participants comprised a total of 120 patients with end-stage renal disease (ESRD) maintained on hemodialysis. The patients were subjected to two groups of treatment-based group divisions: adherent (n = 60) and non-adherent (n = 60). Adherence to treatment for the last 6 months was the criteria for defining dialysis. It was defined as attending at least 90% of all scheduled dialysis sessions over the previous 6 months. Non-adherents were defined as those who missed more than 10% of their sessions in the same period. Adherence to therapy data were collected through self-reports and medical records. Patients were also asked about barriers they face in adhering to treatment, such as financial, psychological, or logistical issues. The inclusion criteria were being aged 18 years or older, diagnosed with ESRD, and being on regular hemodialysis for at least 3 months. Patients with active infections, major surgeries within the last month, or incomplete medical records were excluded from the study.

Collectively collated across the demographic and clinical variables such as age, gender, comorbidities like diabetes, hypertension, and cardiovascular disease, and the duration of dialysis treatment. The following were some of the clinical parameters measured: serum ferritin, transferrin saturation (TSAT), and hemoglobin levels. Serum ferritin was measured by means of a chemiluminescent immunoassay and was defined by a reference range of 30-300 ng/mL. Serum iron/total iron-binding capacity measured TSAT as (serum iron / total iron-binding capacity) × 100, with reference ranges of 20% to 50%. Hemoglobin levels were measured with an automated hematology analyzer, with reference ranges of 13-17 g/dL for males and 12-15 g/dL for females. The variable parameter included serum albumin, parathyroid hormone (PTH), and calcium-phosphorus level records to control for confounding contributions due to CKD progression or nutritional status.

Using descriptive statistics for data analysis, populations were reported to embody the demographic and clinical characteristics of participants. Mean ± standard deviation (SD) or median values with quartile ranges (IQR) were used to show continuous variables. Categorical variables were summarized as the number of observations and proportions. Independent t-tests compared between adherent or non-adherent groups for continuous variables; categorical variables were assessed with chi-square tests. Finally, a multivariate linear regression analysis was used for further analysis on the independent effect of dialysis adherence on the iron status and confounding factors such as age, sex, diabetes, and dialysis duration. All statistical analyses were done using SPSS version 25, with statistical significance levels set at Less than p < 0.05.

Institutional ethical committee was approached for ethical clearance and ethical approval was granted for the study before commencing the study, and all procedures were conducted according to ethical standards ethical clearance ref: ISERC/KTRH/012/24. Written informed consent was obtained from all participants prior to entry into the study. Patients’ identity confidentiality was protected, as data anonymization was applied all along the analysis.

## Results

The study included a total of 120 patients with end-stage renal disease (ESRD) who were on maintenance hemodialysis. The participants were grouped into two groups according to their dialysis adherence; adherent n = 60 and non-adherent n = 60. Below is a summary of demographic and clinical data among the two groups.

### Demographic and Clinical Characteristics

The mean age of the adherent group was 58.2 ± 12.4 years with the non-adherent group having mean age of 60.5 ± 14.7 years (p = 0.35). Gender-wise, no significant difference was observed in the two groups as there were 52% adherents and 48% non-adherent males (p = 0.78). In addition, both groups were also not significantly different in terms of comorbidities like diabetes, hypertension and cardiovascular diseases (p > 0.05 for all). The mean duration of dialysis treatment was 5.4 ± 3.1 years in the adherent patients compared to 5.7 ± 3.3 years in the non-adherent ones (p = 0.620) as illustrated in table 1 below.

**Table 1:**
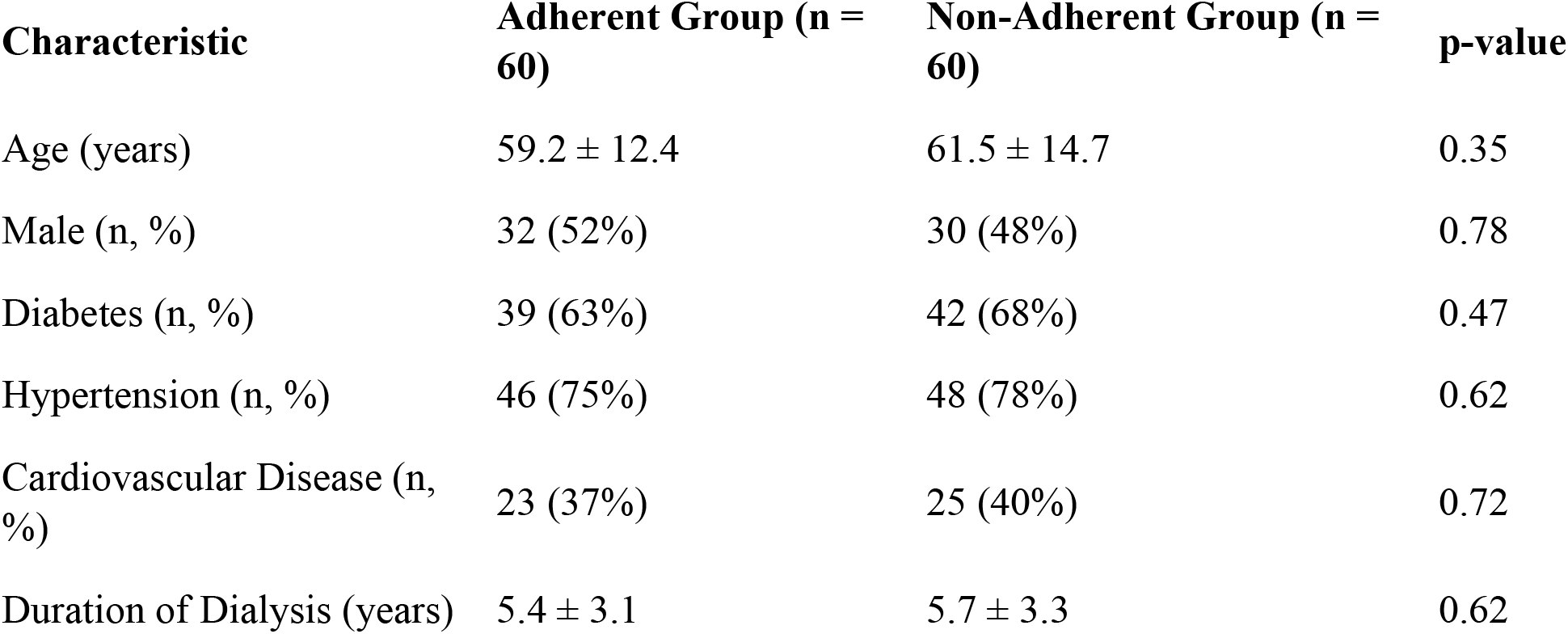
Demographic and Clinical Characteristics of Study Participants.

| Characteristic | Adherent Group (n = 60) | Non-Adherent Group (n = 60) | p-value |
| --- | --- | --- | --- |
| Age (years) | $59.2 \pm 12.4$ | $61.5 \pm 14.7$ | 0.35 |
| Male (n, %) | 32 (52%) | 30 (48%) | 0.78 |
| Diabetes (n, %) | 39 (63%) | 42 (68%) | 0.47 |
| Hypertension (n, %) | 46 (75%) | 48 (78%) | 0.62 |
| Cardiovascular Disease (n, %) | 23 (37%) | 25 (40%) | 0.72 |
| Duration of Dialysis (years) | $5.4 \pm 3.1$ | $5.7 \pm 3.3$ | 0.62 |

### Iron Profile

The iron profile outcomes evidenced that the participants in the adherent group presented a significantly higher serum ferritin level (p = 0.03) and transferrin saturation (TSAT) (p = 0.02) in comparison with their counterparts in the non-adherent group. Hemoglobin levels were also significantly higher in the adherent group (p = 0.04). There seems to be a better iron status and improved anemia control in those who adhered to the dialysis regimen as compared to the ones who did not, as illustrated in table 2 below.

**Table 2:** Iron Profile Parameters of Study Participants.

| Parameter | Adherent Group (n = 60) | Non-Adherent Group (n = 60) | p-value |
| --- | --- | --- | --- |
| Serum Ferritin (ng/mL) | $236.6 \pm 120.2$ | $186.2 \pm 105.3$ | 0.03 |
| Transferrin Saturation (%) | $33.4 \pm 9.3$ | $28.8 \pm 10.2$ | 0.02 |
| Hemoglobin (g/dL) | $12.5 \pm 1.8$ | $11.2 \pm 2.1$ | 0.04 |
| Serum Iron ($\mu\text{g/dL}$) | $84.2 \pm 25.6$ | $76.8 \pm 22.9$ | 0.12 |

### Multivariate Analysis

A multivariate linear regression analysis was done to evaluate the independent effect of dialyzing adherence on iron profile variables while adjusting for potential confounders such as age, sex, diabetes, and duration of dialysis. The results showed that adherence to dialysis significantly remained as independent predictor of increased serum ferritin (β = 0.45, p = 0.02), higher TSAT (β = 0.37, p = 0.03), and larger hemoglobin levels (β = 0.29, p = 0.04) even after controlling for the covariates mentioned earlier. Non-adherence was significantly identified risk factor leading to poorer iron indices in patients with ESRD.

### Association of Adherence and Iron Deficiency

Iron deficiency anemia as defined using serum ferritin <100 ng/mL and TSAT <20% was present among nonadherent patients. Out of the 60 patients who did not adhere, 38 (63%) had iron deficiency anemia, while 24 (40%) of the adherent group were found to be afflicted (p = 0.01). This thus strengthens the evidence that nonadherent patients spend more time on dialysis than those who are nonadherent due to higher iron deficiency and risk of anemia.

## Discussion

Some analyses provide evidence that adherence to dialysis is positively related to the improved iron profile of CKD patients undergoing hemodialysis. More notably, serum ferritin, transferrin saturation (TSAT), and hemoglobin levels were found to be much higher in adherent patients than in non-adherent ones. These results align well with the increasing body of evidence highlighting the adverse impact of non-adherence to dialysis on iron metabolism and anemia management in hemodialysis patients.

Iron deficiency is a common problem among dialysis patients and contributes very significantly to the burden of anemia, one of the most common co-morbid conditions accompanying CKD. Non-adhered patients have poor iron status associated with poor iron availability and measured by lower serum ferritin and TSAT levels. Previous studies found the same results concerning poor adherence to dialysis programs, being a major contributor to iron deficiency anemia in patients with ESRD [9][10]. Deviation from that effectively leads to missed dates of dialysis sessions, incomplete waste product removal and overhydration, and an underperforming erythropoiesis, which all degrade iron metabolism and amplify anemia to some extent.

Several mechanisms allow for the association to be explained whereby adherence to dialysis and iron profiles is related. For one, such patients have a higher likelihood of receiving an uninterrupted and complete course of erythropoiesis-stimulating agents (ESAs) and iron supplementation than their non-adherent counterparts; this means that very beneficial conditions should be created for hemoglobin levels in their patients, particularly patients with ESRD. In several studies, adherence to ESA therapy and the intravenous form of iron treatment impacts positively on iron parameters and better control of the anemia [11]. On the other hand, for the non-adherent patients, there are missed doses of such vital medications.

The study found that the prevalence of iron deficiency anemia was significantly higher in non-adherent patients, that is, 63% of patients had serum ferritin <100 ng/mL and TSAT <20% in comparison to the just 40% of patients with adherence. This corroborates the report by Kalantar-Zadeh et al. where patients with poor dialysis adherence are prone to develop iron deficiency anemia as a result of poor iron supplementation and interrupted treatment regimens [13]. Moreover, iron deficiency anemia in non-adherent patients will also put them in a vicious cycle where untreated anemia will further worsen patient fatigue, reduce physical functionality, and hence quality of life, hindering treatment adherence.

Interestingly, our study showed significant differences between both groups in serum ferritin and TSAT but not with regard to serum iron level, which was not statistically significant (p = 0.12). This is perhaps due to the complex nature of iron metabolism in dialysis patients in which inflammation, malnutrition, and comorbidity can also affect serum iron levels independently of adherence [14] (15). The reason for this is that serum iron provides only a snapshot of the available iron at any given instant and lacks any measurement of iron stores or the efficiency of iron utilization by the bone marrow, which would be better suited in the explanation of the observed differences in ferritin and TSAT.

The multivariate regression analysis conducted in our study strengthens the premise that dialysis adherence alone is an independent predictor of improved iron status. This evidence remains intact with the account of confounding factors age, sex, diabetes, and duration of dialysis. It is consistent with many earlier studies that reported a significant role of dialysis treatment in the prevention of complications from iron deficiency and anemia among CKD patients [16][17]. In addition, the findings underscore regular monitoring of iron parameters with relevant interventions targeted to improve adherence in this particular population.

## Conclusion

As per the discussion, the present study adds to the evidence for the importance of dialysis adherence to achieve and maintain favorable iron status for and anemia management in hemodialysis patients with CKD. Improving patient adherence to treatment programs for hemodialysis could then represent a major strategy for optimizing iron metabolism, decreasing iron deficiency anemia, and improving the health conditions of end-stage renal disease patients. Future interventions should examine the barriers to adherence and help eliminate disconnectedness in access to iron and erythropoiesis-stimulating therapies.

## Data availability

The data of the findings of this study are all shared on this article

